# Longitudinal antibody correlates of SARS-CoV-2 infection in a US household cohort during Omicron waves

**DOI:** 10.64898/2026.09.10.26362752

**Authors:** Yangyupei Yang, Amy Callear, Matthew Smith, Casey Juntila-Raymond, Elie-Tino Godonou, Emileigh Johnson, Adam S. Lauring, William J. Fitzsimmons, Jefferson M. Jones, Claire M. Midgley, Arnold S. Monto, Emily T. Martin

## Abstract

**Background:** SARS-CoV-2 vaccines have been updated based on circulating variants and the antibody protection from prior vaccination and infection against emerging variants. We evaluated longitudinal antibody correlates of SARS-CoV-2 infection risk during Omicron waves.

**Methods:** We used data from a longitudinal cohort study from 2021 through 2025 with prospective surveillance for acute respiratory illness, and repeated blood collection. A subset of participants also provided weekly respiratory specimens to identify asymptomatic infections. Serum samples were tested using multiplex assays measuring anti-Spike (S) binding IgG and angiotensin-converting enzyme 2 (ACE2) inhibition (a measure of inhibition of the ACE2 receptor interaction) across multiple SARS-CoV-2 variants. Cox models were used to evaluate associations between antibody, prior vaccination and infection, and SARS-CoV-2 infections.

**Findings:** Among 119 participants who contributed 438.8 person-years of follow-up, 81 symptomatic SARS-CoV-2 infections and 4 asymptomatic infections (from 33 participants with weekly swabs) were detected. Each two-fold increase in anti-S IgG concentration was associated with a 15% lower hazard of symptomatic infection (hazard ratio [HR]=0.85, 95%CI:0.82–0.88). Similarly, each 10% increase in ACE2 inhibition was associated with a 7% lower infection hazard (HR=0.93, 95% CI:0.90–0.95). After adjusting for antibody levels, prior SARS-CoV-2 infection remained protective, while recent vaccination was not associated with infection hazard. The peak predicted antibody after repeat COVID-19 vaccinations was estimated to confer 62–65% protection against symptomatic infection.

**Interpretation:** Higher levels of binding and functional antibodies were associated with reduced the risk of SARS-CoV-2 infections across multiple waves between 2021—2025. These findings support the use of longitudinal serologic surveillance to inform vaccine evaluation and future booster strategies against evolving SARS-CoV-2 variants.

**Research in Context:** *Evidence before this study:* We searched PubMed and Google Scholar for studies published between January 1, 2020 and May 1, 2026 using combinations of the terms “SARS-CoV-2”, “COVID-19”, “correlates of protection”, “antibody correlates”, “Omicron”, and “longitudinal cohort”. Previous studies consistently demonstrated that SARS-CoV-2 antibody levels, particularly neutralizing antibodies, are associated with protection against infection. Most evidence was from vaccine clinical trials, healthcare worker cohorts, or observational studies evaluating a single epidemic wave or variant period. In many studies, antibody measurements were obtained at a single time point, often shortly after vaccination, and related to subsequent infection risk months later. Few studies incorporated repeated antibody measurements over time, and most focused on either binding antibodies or neutralizing antibodies alone. During the Omicron era, correlates of protection studies were frequently limited to specific subvariants such as BA.1 or BA.5, and relatively little evidence exists on how antibody correlates perform across successive waves characterized by rapidly changing variants. To our knowledge, no previous study in the United States has evaluated longitudinal antibody correlates of protection across the multiple Omicron waves using repeated serologic measurements, comprehensive vaccination and infection histories, and both symptomatic and asymptomatic infection surveillance.

*Added value of this study:* This study used data from a prospective longitudinal U.S. household cohort, enabling evaluation of antibody correlates of protection across multiple Omicron waves from January 2022 through January 2025. More than 90% of participants had been enrolled before the start of the COVID-19 pandemic, providing detailed pre-pandemic baseline information and complete longitudinal histories of SARS-CoV-2 vaccination and infection. Unlike most previous studies that relied on a single antibody measurement, we incorporated all available serum specimens (median 5 [IQR 3-8] specimens per participant) and modeled antibody levels as time-varying exposures. We evaluated both spike-binding IgG antibodies and ACE2 binding inhibition, a functional measure closely related to neutralizing activity, and examined multiple composite antibody definitions to assess the robustness of findings. In addition to symptomatic infections identified through active respiratory illness surveillance, a subset of participants underwent weekly respiratory sampling, allowing inclusion of asymptomatic infections. Across all analyses, higher antibody levels were consistently associated with reduced infections, and findings were similar across binding and functional antibody measures and across alternative composite antibody definitions.

*Implications of all the available evidence:* Together with previous studies, these findings support SARS-CoV-2 variant-specific antibody responses as correlates of protection against infection, particularly against dominant circulating variants. The consistency of associations across multiple Omicron waves, evolving variants, and alternative antibody measurements suggests that longitudinal serologic surveillance can provide meaningful information about population susceptibility even in settings with complex vaccination and infection histories. After adjustment for measured antibody levels, recent vaccination was not associated with additional reduction in infection hazard, whereas prior infection remained associated with lower infection hazard. In other words, the protection associated with recent vaccination was largely captured by measured antibody responses, while prior infection was associated with protection beyond that captured by measured antibody levels. These results have implications for vaccine evaluation, interpretation of serosurveys, and future booster vaccine strategies as SARS-CoV-2 continues to evolve.

## Background

COVID-19, caused by SARS-CoV-2, has imposed a significant health burden globally, with the virus undergoing rapid evolution and causing severe diseases^1^. Vaccinations and previous infections induce immune responses that can reduce the risk of developing severe disease and/or future infections^2^. COVID-19 vaccines produce robust antibody responses against the Spike protein after receipt of the primary series, with further increases following additional vaccinations^3^. SARS-CoV-2 infection-induced antibodies can persist months after infection^4,5^. However, research has shown that infection can occur despite the presence of high antibody levels after infection or vaccination^6,7^. Those observations highlight the need to understand the amount of protection that can be attributed to antibodies and determine how antibody-derived protection may have changed in the recent years with circulating Omicron subvariants. Interpretation of antibody protection against pre-specified endpoints, such as hospitalization, symptomatic or asymptomatic infection, has been challenging due to multiple factors, including differences in assay platforms used to measure binding or functional SARS-CoV-2 antibodies, and the continual emergence of new variants (or subvariants) with immune escape potential. Furthermore, existing longitudinal studies on SARS-CoV-2 antibodies are often limited by short study durations, low sampling frequency, or infrequent follow-up^8–15^. No previous study in the U.S. has followed participants from before the pandemic and across multiple Omicron waves with repeated serologic measurements, limiting our understanding of how antibodies from incident vaccination and infection have evolved over time as participants obtain additional vaccinations or infections.

Despite extensive research during the pandemic, the quantitative antibody levels required to protect against SARS-CoV-2 infection during different Omicron waves remain unclear. Establishing these correlates of protection is important for interpreting serologic data, evaluating vaccine performance, and guiding vaccine strategies as the virus continues to evolve. To address these gaps, we conducted a longitudinal analysis using data from the Household Influenza Vaccine Evaluation (HIVE), a household cohort in Southeast Michigan with repeated serologic measurements and infection surveillance spanning multiple Omicron waves^16^. This design provides a unique opportunity to quantify how SARS-CoV-2 antibody levels relate to subsequent infection hazards across multiple Omicron waves over an extended period of viral circulation.

## Methods

### 2.1 Study population

We used data from the HIVE study, an ongoing prospective household cohort that has conducted active acute respiratory illness (ARI) surveillance since 2011^16^. Participants were instructed to report illnesses meeting a standard case definition: the onset of 2 or more ARI symptoms from anyone in the household, including cough, fever/feverishness, nasal congestion, chills, headache, body aches, and/or sore throat (Supplementary methods). During the SARS-CoV-2 pandemic, participants reporting two or more ARI symptoms provided nasal or throat swabs for detection of SARS-CoV-2 and other respiratory viruses using molecular testing, and serum specimens were collected twice yearly, and before and after influenza or COVID-19 vaccination.

A subset of HIVE participants collected weekly nasal swabs regardless of symptoms from September 2022 through July 2024, enabling identification of both symptomatic and asymptomatic SARS-CoV-2 infections^17^; nasal swabs were tested for SARS-CoV-2 using multiplex reverse transcriptase-polymerase chain reaction (RT-PCR)^16^.

Each participant, or their parent or legal guardian, completed an enrollment survey detailing demographics, household characteristics, chronic medical conditions, COVID-19 vaccination history, and prior SARS-CoV-2 infection. Participants were surveyed annually to update demographic information. COVID-19 vaccination status was additionally verified using state immunization information systems and electronic medical records, while chronic medical conditions were obtained from self-report and electronic medical records.

This study was reviewed and approved by the University of Michigan Institutional Review Board, and was conducted consistent with applicable federal law and CDC policy (See 45 C.F.R. part 46.114; 21 C.F.R. part 56.114). All household members ≥18 years of age provided written informed consent, and written parental consent was provided for minors.

Participants were eligible for this analysis if they were active HIVE cohort participants and had at least one serum specimen collected during July 1 and December 31, 2021. These criteria ensured availability of a pre-Omicron antibody measurement for longitudinal follow-up. Participants vaccinated before January 1, 2022 were required to have at least one serum specimen within 30 days post-vaccination, while participants not vaccinated before January 1, 2022 were required to have at least one serum specimen collected during July 1, 2021 and January 1, 2022. Although HIVE enrollment began before the COVID-19 pandemic and historical vaccination and infection data were available, this analysis focused on SARS-CoV-2 infections occurring during the Omicron period (2021 through 2025).

### 2.2 Serological assays and variant classification

Serum specimens from eligible participants were tested for IgG antibodies to the SARS-CoV-2 spike protein using multiplex electrochemiluminescence assay (Meso Scale Discovery, Rockville, MD, USA)^18^. An additional angiotensin-converting enzyme 2 (ACE2) assay was used to evaluate the antibody’s capacity to inhibit the binding of ACE2 to various SARS-CoV-2 antigens. To account for changes in circulating variants over time, we defined multiple sero-surveillance periods and used different assay panels for each period (Table S1). Each panel included antigens representing SARS-CoV-2 variants and sublineages circulating during the corresponding surveillance period^19^. Serum samples were diluted and processed following the manufacturer’s protocol^20^ and described elsewhere^18^.

SARS-CoV-2 infections were defined only using RT-qPCR-confirmed positive respiratory swab specimens, and serologic evidence was not used to define infections. Nasal swab specimens testing positive for SARS-CoV-2 by RT-qPCR collected within 30 days of a prior positive test were considered the same infection episode, and the infection date was defined as the first positive specimen collection date. Whole-genome sequencing was attempted on all SARS-CoV-2 infection-positive swab specimens with RT-qPCR cycle threshold ≤ 32. Swab specimens were processed for whole-genome sequencing using the ARTIC Network protocol (v5.3.2 primers) on an Illumina instrument. Lineages were assigned using PANGO^17,21^, and were used to identify Omicron variants and define the Omicron waves (Figure 1; Table S2). Omicron wave 1 was defined as January 1 through June 30, 2022, corresponding to predominant BA.1/BA.2 circulation within the cohort; wave 2 was defined as July 1 through December 31, 2022, corresponding to predominant BA.4/BA.5 circulation; and wave 3 was defined as January 1, 2023 through January 31, 2025, corresponding to the period when XBB- and JN-related variants predominated within the HIVE cohort.

**Figure 1:**
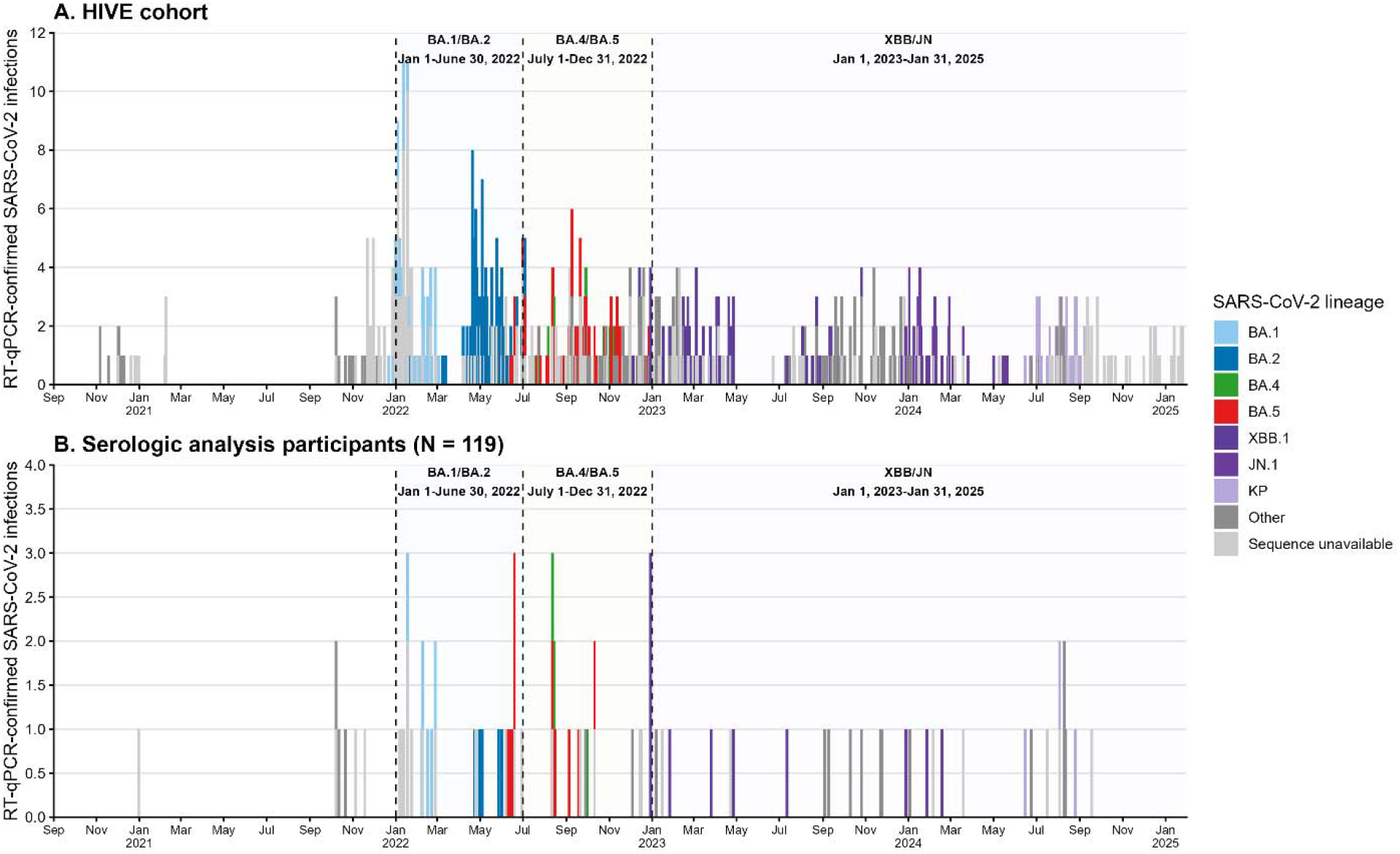
Temporal distribution of RT-qPCR-confirmed SARS-CoV-2 infections and dominant circulating variants during Omicron periods in the HIVE cohort and analysis participants, September 2020–January 2025. Bars represent all RT-qPCR-confirmed SARS-CoV-2 infection episodes identified through (A) HIVE surveillance, and (B) 119 participants included in this serologic analysis. Colors indicate SARS-CoV-2 lineages assigned by whole genome sequencing. “Other” includes successfully sequenced SARS-CoV-2 lineages that were detected but not included among the major subvariant categories shown separately (e.g., BQ.1.1). Vertical dashed lines indicate the study-defined Omicron wave periods used in subsequent analyses: Wave 1 (January 1–June 30, 2022), Wave 2 (July 1–December 31, 2022), and Wave 3 (January 1, 2023–January 31, 2025). Wave definitions were based on the predominant SARS-CoV-2 variants circulating in Michigan and supported by sequencing data from infections detected in the HIVE cohort. Distinct infection episodes were defined as RT-qPCR-positive respiratory specimens collected more than 14 days apart, with the infection date assigned as the date of the first positive specimen.

### 2.5 Statistical analysis

Descriptive statistics were used to compare demographic information and household characteristics of participants for categorical variables and median (IQR) for continuous variables. P values were calculated using χ2 tests for categorical variables and t-tests for continuous variables at the 0.05 significance level. IgG Geometric Mean Concentrations (GMCs) and mean ACE2 percent inhibition were calculated for each Omicron wave as defined above.

To account for the high correlation in detected antibody levels observed among antigens included in the multiplex assay panels^18^, we developed several composite variables to generate a single summary for IgG concentration and for ACE2 percent inhibition of each serum specimen, rather than evaluating each antigen-specific response separately. These composite measures served as primary exposures in our analyses. The first composite variable approach utilized the antigen of the predominant circulating Omicron variant during each wave, defined as the variant with the highest circulation proportion during the corresponding period among HIVE participants: BA.2.75 for wave 1, BA.5 for wave 2, XBB.1.5 during the early period of wave 3 (January 2023–December 2023), and JN.1 during the later period of wave 3 (January 2024–January 2025). Additional composite variables were developed for use in sensitivity analyses and were defined as the mean, minimum, and maximum antibody responses across all antigen targets included in each testing panel (Table S2). We calculated the IgG GMCs and the mean ACE2 percent inhibition across all tested antigens. We also identified the maximum and minimum values for both IgG concentration and ACE2 inhibition to capture the range of immune responses within each sample.

We estimated the IgG antibody concentration and ACE2 percent inhibition following each dose of COVID-19 vaccination or infection. Serum samples were included if collected between 3 and 180 days after a vaccination or infection event. Quadratic mixed-effects models were fitted to characterize antibody kinetics, with fixed effects for vaccine dose or infection event and random intercepts for individuals (Supplementary Methods). Predicted mean IgG concentration and ACE2 percent inhibition with corresponding 95% confidence intervals were derived to describe antibody responses following vaccination or infection over time. In sensitivity analyses focused on peak post-exposure responses, analyses were repeated restricting serum specimens to those collected between 14 and 60 days after vaccination; analyses were not powered to assess responses 14 to 60 days after infection.

We estimated the association between antibody levels and time to SARS-CoV-2 infection using the Andersen–Gill extension of the Cox proportional hazards model. This approach allows modeling of repeat infection events in a given participant while incorporating antibody composite measures of all available serum specimens as time-varying covariates, adjusting for confounding by age (Supplementary Methods). To address within-household correlation, we included a shared frailty term to account for dependence among individuals within the same household. Analyses initially focused on symptomatic SARS-CoV-2 infections identified through ARI surveillance in the HIVE cohort and were subsequently expanded to include asymptomatic infections among participants with weekly swabs. We conducted sensitivity analyses to evaluate the robustness of the primary results. These analyses examined alternative definitions of composite antibody measures, different assumptions for the infection risk period, and alternative model specifications for vaccination and prior infection covariates (Supplementary Methods).

To visualize antibody dynamics and their relationship with infection risk, we additionally fit generalized additive mixed models (GAMMs) to estimate smooth trends in IgG antibody concentration and ACE2 percent inhibition over time. These models incorporated smooth functions to flexibly capture nonlinear antibody trajectories and included random intercepts for individuals to account for repeated measurements within participants (Supplementary Methods).

All statistical analyses were conducted using R (version 4.3.1).

## 2 Results

### 3.1 Participant and household characteristics

A total of 119 participants (of 986 HIVE participants) from 66 households were included in this analysis (Table 1, Figure S1). Among those included, 107 (90%) participants had begun enrollment in the HIVE study before the COVID-19 pandemic. The median age of included participants was 40 years (range: 1-83 years), with 68% being adults over 18 years. The age distribution of included participants differed from the overall HIVE cohort, which had a median age of 18 years (range: 0-83 years). Of the included participants, 60% were female, and 71% were non-Hispanic white. Around 36% of participants reported at least one comorbidity (note, only 23% reported a comorbidity in the overall HIVE cohort), while 34% children attended daycare or school during the pandemic (note, 50% was observed among all HIVE participants). The median number of adults in a participant’s household was 2 (range: 1-7), and the median number of children was also 2 (range: 0-7).

**Table 1:**
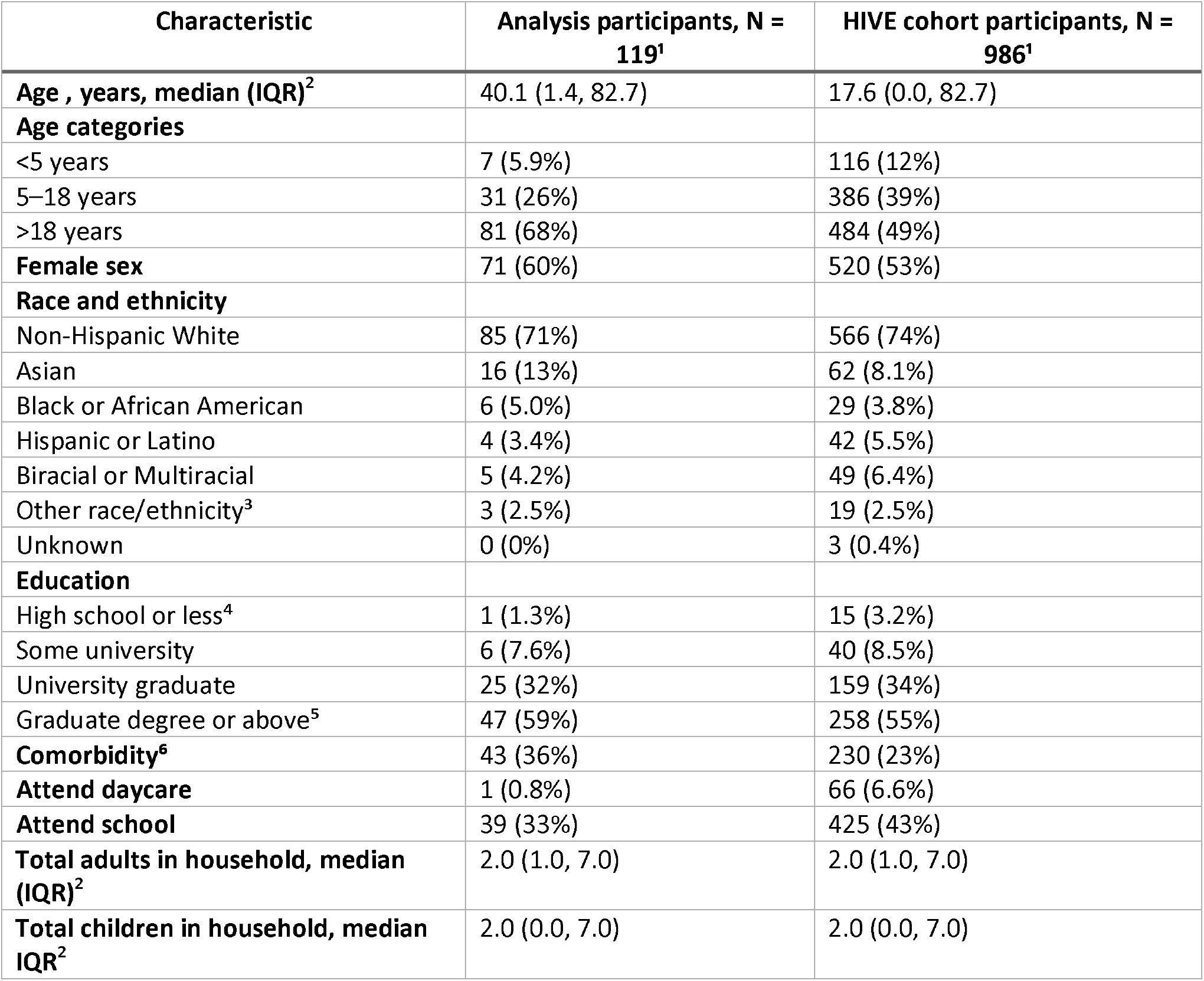
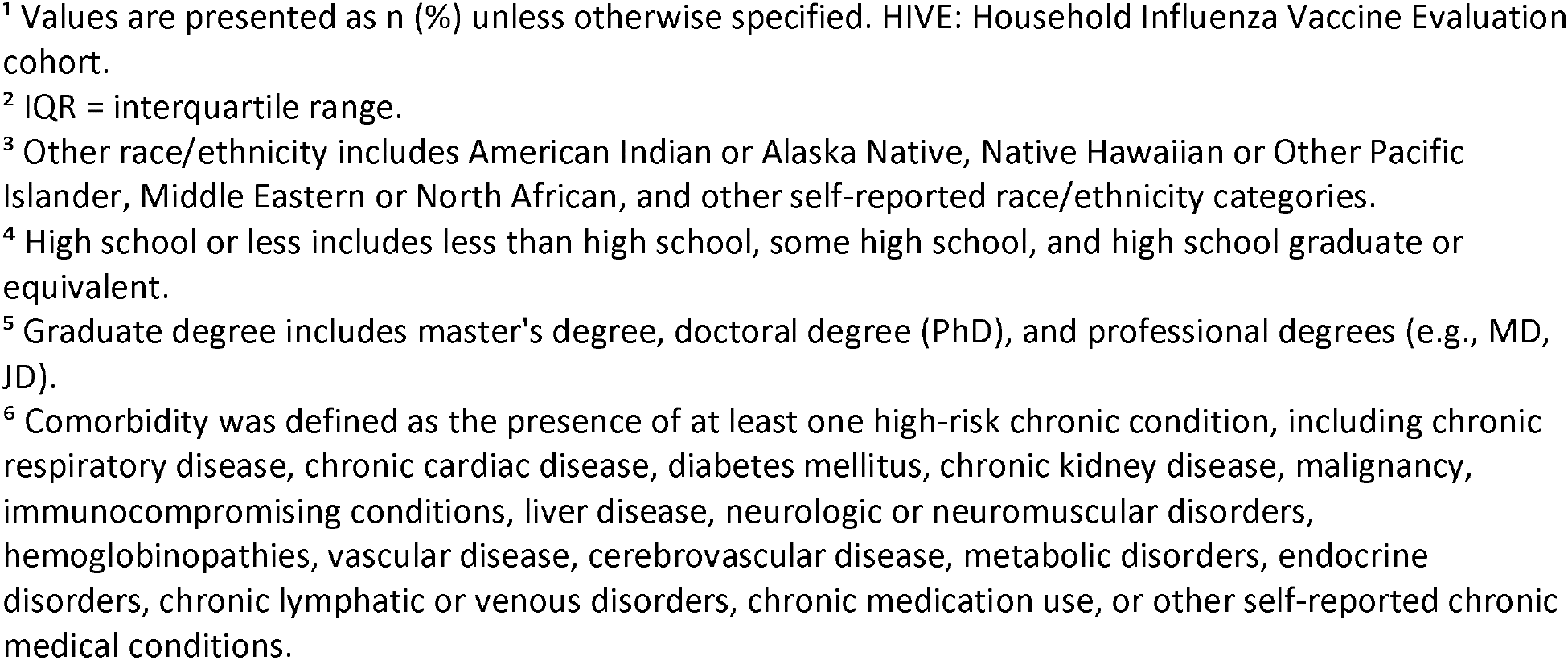
Participant and household characteristics of participants included in this SARS-CoV-2 antibody correlates of protection analysis and the overall HIVE cohort (2021—2025).

| Characteristic | Analysis participants, N = 119<sup>1</sup> | HIVE cohort participants, N = 986<sup>1</sup> |
| --- | --- | --- |
| **Age , years, median (IQR)<sup>2</sup>** | 40.1 (1.4, 82.7) | 17.6 (0.0, 82.7) |
| **Age categories** |  |  |
| <5 years | 7 (5.9%) | 116 (12%) |
| 5–18 years | 31 (26%) | 386 (39%) |
| >18 years | 81 (68%) | 484 (49%) |
| **Female sex** | 71 (60%) | 520 (53%) |
| **Race and ethnicity** |  |  |
| Non-Hispanic White | 85 (71%) | 566 (74%) |
| Asian | 16 (13%) | 62 (8.1%) |
| Black or African American | 6 (5.0%) | 29 (3.8%) |
| Hispanic or Latino | 4 (3.4%) | 42 (5.5%) |
| Biracial or Multiracial | 5 (4.2%) | 49 (6.4%) |
| Other race/ethnicity<sup>3</sup> | 3 (2.5%) | 19 (2.5%) |
| Unknown | 0 (0%) | 3 (0.4%) |
| **Education** |  |  |
| High school or less<sup>4</sup> | 1 (1.3%) | 15 (3.2%) |
| Some university | 6 (7.6%) | 40 (8.5%) |
| University graduate | 25 (32%) | 159 (34%) |
| Graduate degree or above<sup>5</sup> | 47 (59%) | 258 (55%) |
| **Comorbidity<sup>6</sup>** | 43 (36%) | 230 (23%) |
| **Attend daycare** | 1 (0.8%) | 66 (6.6%) |
| **Attend school** | 39 (33%) | 425 (43%) |
| **Total adults in household, median (IQR)<sup>2</sup>** | 2.0 (1.0, 7.0) | 2.0 (1.0, 7.0) |
| **Total children in household, median IQR<sup>2</sup>** | 2.0 (0.0, 7.0) | 2.0 (0.0, 7.0) |

<sup>1</sup> Values are presented as n (%) unless otherwise specified. HIVE: Household Influenza Vaccine Evaluation cohort.
<sup>2</sup> IQR = interquartile range.
<sup>3</sup> Other race/ethnicity includes American Indian or Alaska Native, Native Hawaiian or Other Pacific Islander, Middle Eastern or North African, and other self-reported race/ethnicity categories.
<sup>4</sup> High school or less includes less than high school, some high school, and high school graduate or equivalent.
<sup>5</sup> Graduate degree includes master's degree, doctoral degree (PhD), and professional degrees (e.g., MD, JD).
<sup>6</sup> Comorbidity was defined as the presence of at least one high-risk chronic condition, including chronic respiratory disease, chronic cardiac disease, diabetes mellitus, chronic kidney disease, malignancy, immunocompromising conditions, liver disease, neurologic or neuromuscular disorders, hemoglobinopathies, vascular disease, cerebrovascular disease, metabolic disorders, endocrine disorders, chronic lymphatic or venous disorders, chronic medication use, or other self-reported chronic medical conditions.

### 3.2 SARS-CoV-2 infections, variants and Omicron waves

Among the 119 included participants, 96 (81%) received at least one COVID-19 vaccine dose by January 31, 2025, including 98% of adults, 48% of children aged 5–18 years, and 29% of children aged <5 years (Table 2). During the study period, 65 (55%) participants experienced at least one RT-qPCR-confirmed symptomatic SARS-CoV-2 infection. A total of 81 symptomatic SARS-CoV-2 infection events were identified through HIVE acute respiratory illness surveillance (Figure 1). Of these, 7 infection events occurred before Omicron circulation, while 17, 26, and 31 participants experienced at least one infection during Omicron waves 1, 2, and 3, respectively. Wave 1 was characterized predominantly by BA.1/BA.2 infections, wave 2 by BA.4/BA.5 infections, and wave 3 by XBB- and JN-related infections. The total number of infection events exceeded the number of infected participants because reinfections occurred during follow-up. Fourteen participants experienced multiple RT-qPCR-confirmed infection episodes, including 12 participants with two infections and 2 participants with three infections. Distinct infection episodes were defined as RT-qPCR-positive respiratory specimens collected more than 30 days apart.

**Table 2:** COVID-19 vaccination and symptomatic SARS-CoV-2 infection history among analysis participants by age group, HIVE cohort (2021—2025).

| Characteristic | <5 years<br>N = 7<sup>1</sup> | 5-18 years<br>N = 31<sup>1</sup> | >18 years<br>N = 81<sup>1</sup> | Total<br>N = 119<sup>1</sup> | p-value<sup>2</sup> |
| --- | --- | --- | --- | --- | --- |
| Ever receive COVID-19 vaccine | 2 (29%) | 15 (48%) | 79 (98%) | 96 (81%) | <0.001 |
| Total COVID-19 vaccine dose by Jan 2025 | 0 (0, 3) | 0 (0, 6) | 6 (0, 9) | 5 (0, 9) |  |
| At least one SARS-CoV-2 infection during study<sup>3,4,5</sup> | 3 (43%) | 14 (45%) | 48 (59%) | 65 (55%) | 0.53 |
| Median total SARS-CoV-2 infections per participants<sup>3,4,6</sup> | 0 (0, 2) | 0 (0, 3) | 1 (0, 2) | 1 (0, 3) |  |
| SARS-CoV-2 infection before Omicron<sup>3</sup> | 0 (0%) | 1 (3.2%) | 6 (7.4%) | 7 (5.9%) | 0.76 |
| SARS-CoV-2 infection during Omicron wave 1<sup>3,4,7</sup> | 0 (0%) | 7 (23%) | 10 (12%) | 17 (14%) | 0.37 |
| SARS-CoV-2 infection during Omicron wave 2<sup>3,4,7</sup> | 3 (43%) | 3 (9.7%) | 20 (25%) | 26 (22%) | 0.18 |
| SARS-CoV-2 infection during Omicron wave 3<sup>3,4,7</sup> | 2 (29%) | 7 (23%) | 22 (27%) | 31 (26%) | 0.97 |

<sup>1</sup> Values are presented as n (column %) or median (range), unless otherwise specified.
<sup>2</sup> P values were calculated using Pearson's $\chi^2$ tests for categorical variables.
<sup>3</sup> SARS-CoV-2 infections presented in this table represent symptomatic RT-qPCR-confirmed infections identified through HIVE acute respiratory illness surveillance. Asymptomatic infections identified through weekly swab surveillance in the subset are not included. Distinct infection episodes were defined as RT-qPCR-positive respiratory specimens collected more than 30 days apart.
<sup>4</sup> Participants could contribute infections to more than one Omicron wave; therefore, counts across wave-specific categories do not equal to the total number of participants infected.
<sup>5</sup> "At least one SARS-CoV-2 infection during study" indicates participants with at least one symptomatic RT-qPCR-confirmed SARS-CoV-2 infection during follow-up.
<sup>6</sup> "Total SARS-CoV-2 infections" represents the total number of symptomatic infection episodes experienced by each participant during follow-up and is summarized as the median (range).
<sup>7</sup> Omicron wave 1 was defined as January 1 to June 30, 2022 (predominantly BA.1/BA.2 circulation), wave 2 as July 1 to December 31, 2022 (predominantly BA.4/BA.5 circulation), and wave 3 as January 1, 2023 to January 31, 2025 (predominantly XBB- and JN-related circulation).

Among the 119 included participants, 33 individuals contributed weekly nasal swabs, enabling detection of both symptomatic and asymptomatic SARS-CoV-2 infections. Among those 33 individuals, during September 2022 through July 2024, an additional 4 asymptomatic infections were detected through weekly swab testing (Figure S1).

### 3.3 Antibody levels across the waves

A total of 663 serum specimens were included in the analysis, with participants contributing a median of 5 (IQR: 3–8) serum specimens per participant. SARS-CoV-2 IgG antibody concentrations increased substantially from pre-Omicron to Omicron waves (Figure S2, Table S3). For the target composite variable, the GMC before Omicron circulation was 3,410.6 AU/mL (95% CI: 1,824.2–6,376.6). During the BA.1/BA.2-dominant wave in early 2022, the GMC increased to 64,701.7 AU/mL (95% CI: 49,569.4–84,453.6), coinciding with vaccination introduction. Antibody levels remained higher than pre-Omicron levels during the BA.4/BA.5-dominant wave in late 2022 and the XBB- and JN-dominant waves in 2023–2025. A similar pattern was observed for the target composite ACE2 percent inhibition, which increased from 13.9% (95% CI: 9.9–17.9%) before Omicron circulation to greater than 40% during the BA.1/BA.2-, BA.4/BA.5-, XBB/JN-dominant waves (Figure S2, Table S4). Trends across other composite antibody measures and variant-specific antigens were consistent with these observations.

### 3.3 Immunogenicity of vaccination and infection

Among included participants, 66 (55%) provided at least one post-vaccination serum sample collected during day 3–180. Results from the quadratic mixed model indicated that SARS-CoV-2 IgG concentrations increased with the number of vaccine doses, with a concentration of 158,117 AU/mL (95% CI: 115,520–216,419) after six doses; beyond 6 doses, data were too sparse to reliably infer a trend (Figure 2, Table S5). A similar immunogenicity pattern was observed for ACE2 percent inhibition. The increase in ACE2 inhibition was steep from one to six vaccine doses, reaching 72.6% (95% CI:65.1– 80.0%) at six doses (Figure 2, Table S6). Sensitivity analyses using alternative composite antibody definitions (Figures S3) and restricting serum specimens to 14–60 days after vaccination (data not shown) showed similar patterns.

**Figure 2:**
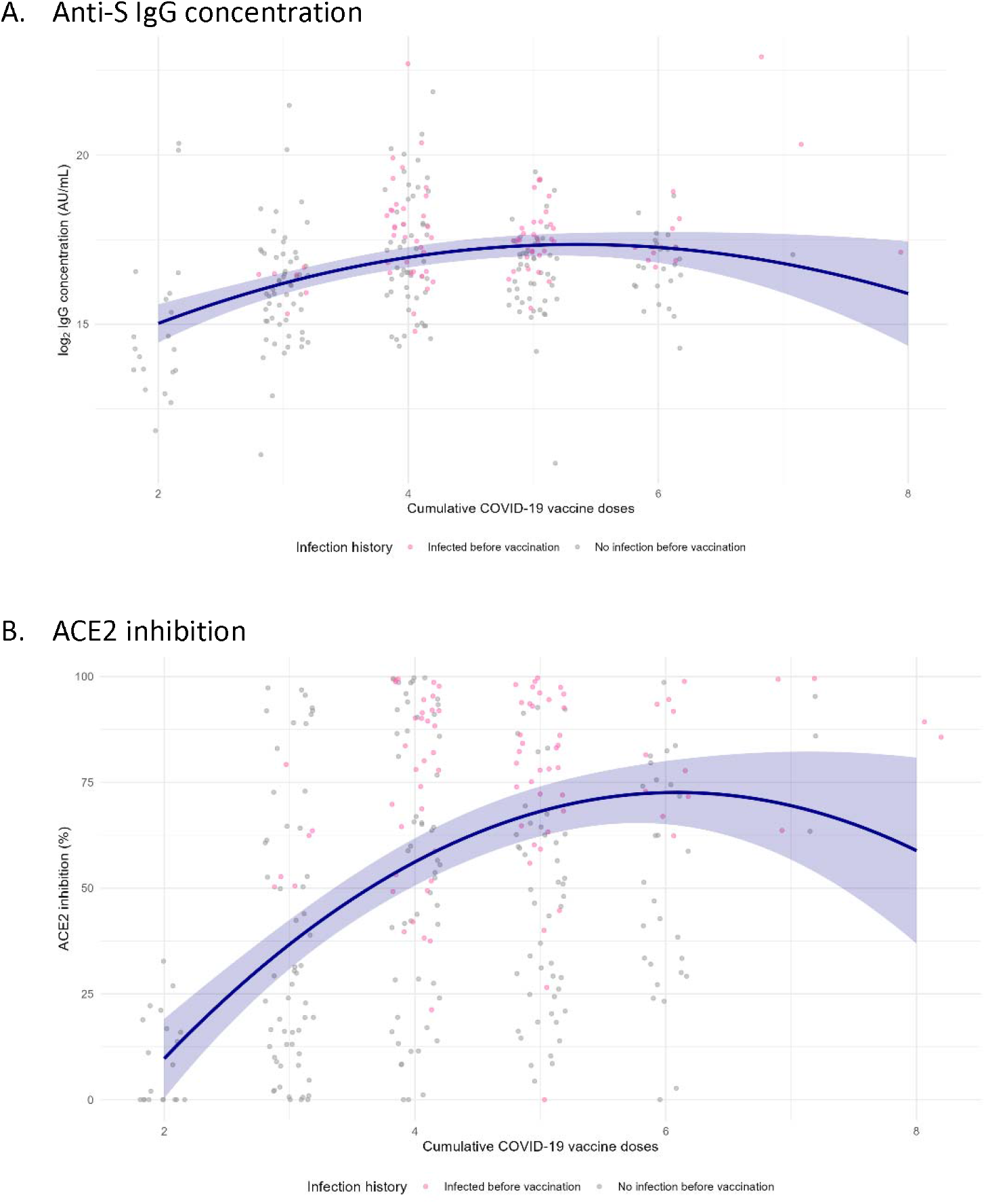
SARS-CoV-2 antibody responses following COVID-19 vaccination, HIVE cohort (2021—2025). Measured levels and predicted trajectories of anti-spike IgG antibody concentration (A) and ACE2 percent inhibition (B) following COVID-19 vaccination, among 66 participants who contributed at least one serum specimen collected between 3 and 180 days after vaccination. Measured antibody levels (dots) are shown by the cumulative number of vaccine doses received, and by prior infection status at the time of vaccination; pink dots represent participants with SARS-CoV-2 infection before each vaccination and gray dots represent participants without SARS-CoV-2 infection before each vaccination. Presented are the primary composite antibody measurements, which used a wave-specific antigen selected to represent the predominant circulating SARS-CoV-2 variant during each analysis period. Predicted trajectories (purple lines) were estimated using quadratic mixed-effects models with fixed effects for vaccine dose and random intercepts for individuals to account for repeated measurements. Shaded bands represent 95% confidence intervals around the predicted mean responses.

Among included participants, 23 (19%) provided at least one post-infection serum sample collected between day 3 and 180. The median time from infection to serum collection was 50 days (IQR: 18–67) following first infections and 80 days (IQR: 62–83) following second infections. Following a first infection, the IgG GMC was 72,793.6 AU/mL (95% CI: 39,756.4–133,284.3) and mean ACE2 inhibition was 51.2% (95% CI: 36.8–65.5%). Following a second infection, the corresponding values were 59,535.1 AU/mL (95% CI: 27,122.8–130,680.8) and 42.4% (95% CI: 17.5–67.2%), respectively. IgG concentrations after infection were substantially higher among individuals who had received at least one vaccine dose prior to infection compared to those who had never been vaccinated (Figure 3, Figure S4, Tables S7-S8). Because of the limited number of post-infection specimens, heterogeneity in vaccination histories, and differences in the timing of specimen collection, formal comparisons between first and second infections were not performed. There were not sufficient data to fit a quadratic mixed model for this subset.

**Figure 3:**
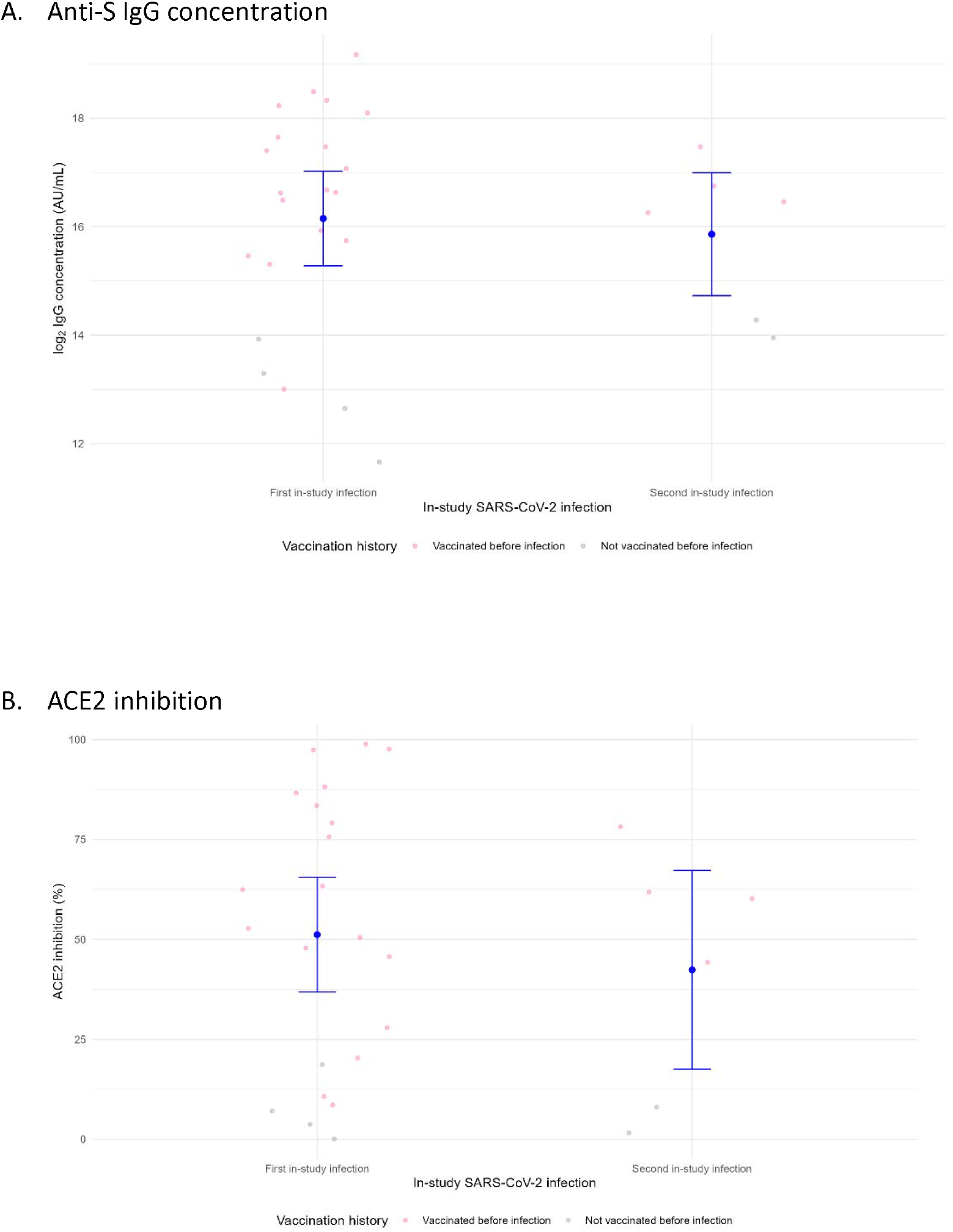
Antibody responses following by first and second in-study Infection, HIVE cohort (2021— 2025). Observed anti-S IgG antibody concentrations (A) and ACE2 percent inhibition (B) among 23 participants contributing 32 post-infection serum specimens collected between 3 and 180 days after RT-qPCR-confirmed SARS-CoV-2 infection. First and second infections refer to the first and second RT-qPCR-confirmed SARS-CoV-2 infection episodes observed during study follow-up; therefore, all participants contributing data following a second infection had previously contributed data following a first in-study infection. Points represent individual serum measurements, and summary estimates are shown with 95% confidence intervals. Due to the limited number of participants and post-infection serum specimens, quadratic mixed-effects models were not fitted for this subset.

### 3.4 IgG antibody concentration and subsequent infection hazard during Omicron

Higher SARS-CoV-2 IgG concentrations were associated with a reduced hazard of symptomatic infection. A two-fold increase in IgG concentration corresponded to a hazard ratio of 0.85 (95% CI: 0.82– 0.88, p<0.001) across all waves combined (Table 3). After including both symptomatic and asymptomatic infections, a two-fold increase in IgG concentration corresponded to a hazard ratio of 0.96 (95% CI: 0.92–0.99, p=0.008) for all infections (Table S9). Alternative composite antibody variables resulted in similar patterns (Table S10).

**Table 3:** Age-adjusted associations of antibody levels, vaccination, and prior SARS-CoV-2 infection with the hazard of symptomatic SARS-CoV-2 infections during Omicron waves, HIVE cohort (2021—2025).

| Exposure | Adjusted Hazard Ratio (95% CI)<sup>1</sup> | P value |
| --- | --- | --- |
| Anti-S IgG concentration (per 2-fold increase)<sup>2</sup> | 0.85 (0.82–0.88) | <0.001 |
| ACE2 inhibition (per 10% increase)<sup>2</sup> | 0.93 (0.90–0.95) | <0.001 |
| Recent vaccination within 180 days<sup>3</sup> | 1.07 (0.93–1.25) | 0.34 |
| Prior SARS-CoV-2 infection<sup>4</sup> | 0.58 (0.45–0.73) | <0.001 |

<sup>1</sup> Adjusted hazard ratios (aHRs) were estimated using separate Andersen–Gill Cox proportional hazards models with a household-level shared frailty term and adjustment for age. Each exposure shown in the table was evaluated in an independent model and was not adjusted for the other exposures listed.
<sup>2</sup> IgG concentration and ACE2 inhibition were derived from the primary composite antibody measure ("target antigen by period"), defined using antigens representing the predominant circulating SARS-CoV-2 variant during each Omicron wave. IgG concentration was modeled on the log<sub>2</sub> scale; hazard ratios represent the change in infection hazard associated with a two-fold increase in antibody concentration. ACE2 inhibition was modeled per 10 percentage-point increase.
<sup>3</sup> Recent vaccination was defined as receipt of a COVID-19 vaccine dose within the previous 180 days. The reported aHR represents the association between recent vaccination and infection hazard after adjustment for age only and should not be interpreted as the effect of vaccination after accounting for antibody levels.
<sup>4</sup> Prior infection was defined as a previous RT-qPCR-confirmed SARS-CoV-2 infection occurring at least 30 days before the current infection event. The reported aHR represents the association between prior infection and infection hazard after adjustment for age only and should not be interpreted as the effect of prior infection after accounting for antibody levels.

Across the observed range of IgG concentrations, higher antibody levels were associated with an increased probability of protection. Among our study participants, GAMM-predicted infection risk decreased as IgG concentrations increased. The IgG concentrations corresponding to predicted infection probabilities of 50% and 30%, respectively (equivalent to 50% and 70% predicted protection), were approximately 15,880 AU/mL and 955,194 AU/mL (Figure 4). The peak IgG concentration predicted by the immunogenicity model (158,117 AU/mL) was associated with an estimated infection probability of 37.7% (95% CI: 35.3%–40.2%), corresponding to approximately 62.3% predicted protection against all infections.

**Figure 4:**
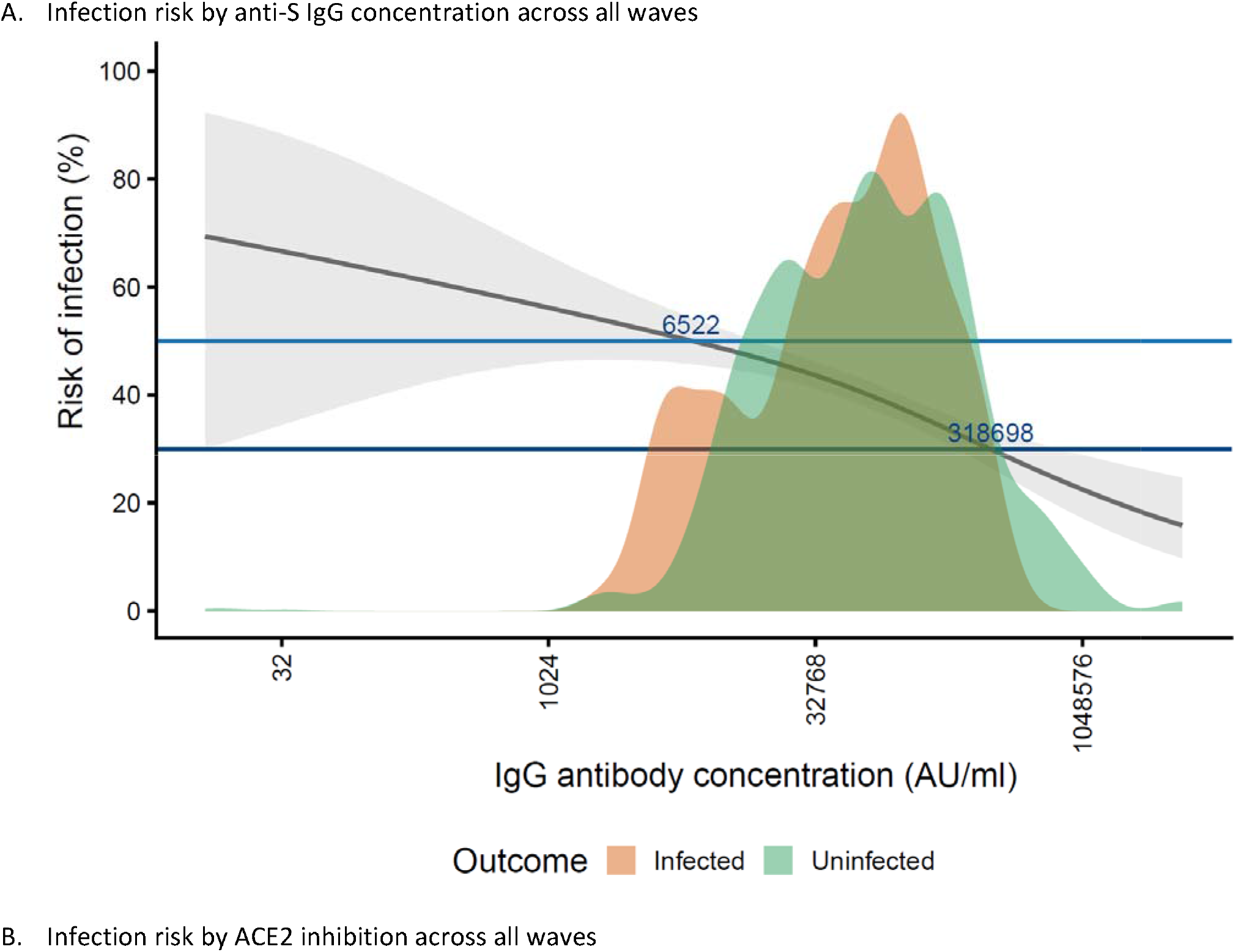
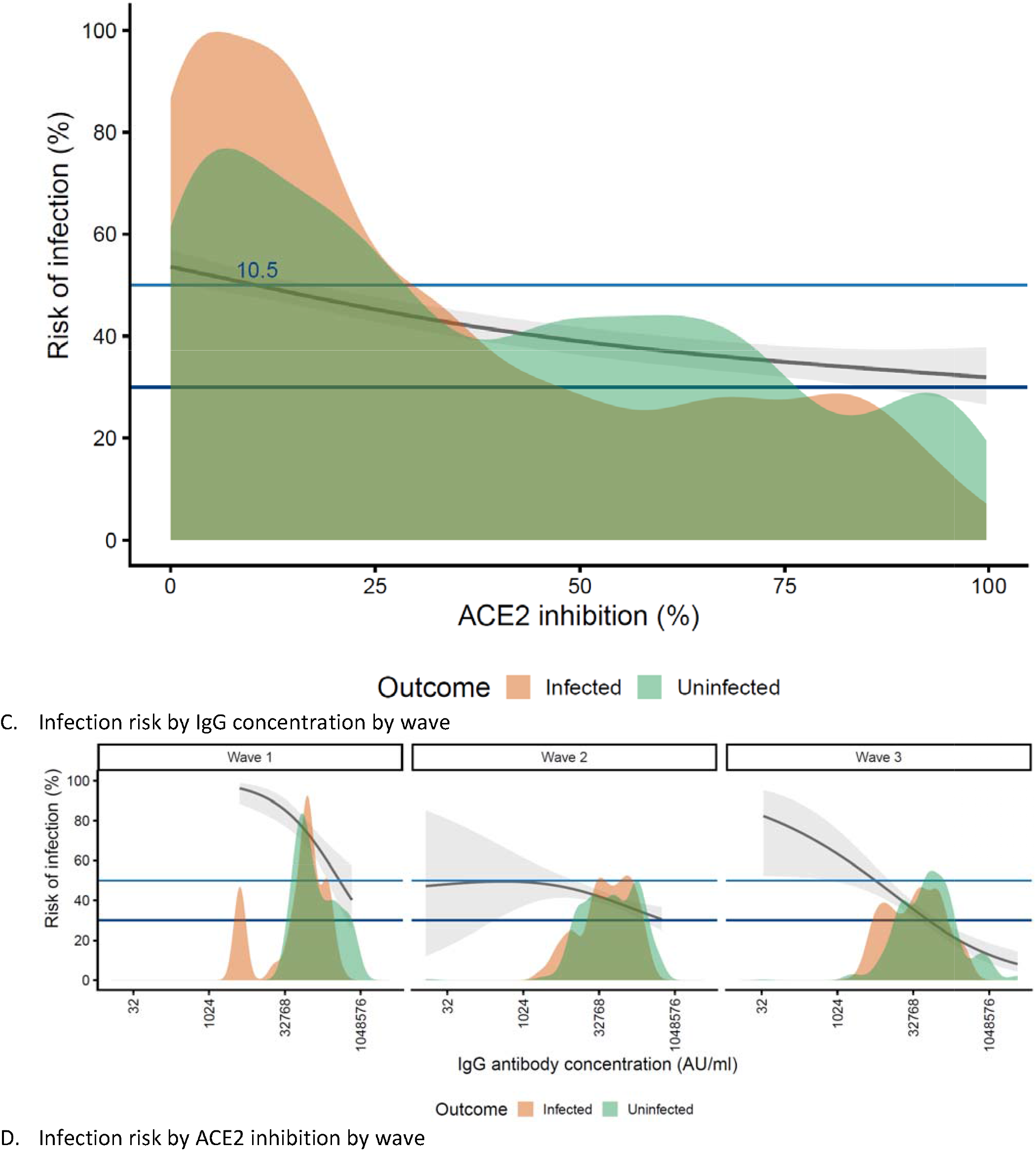
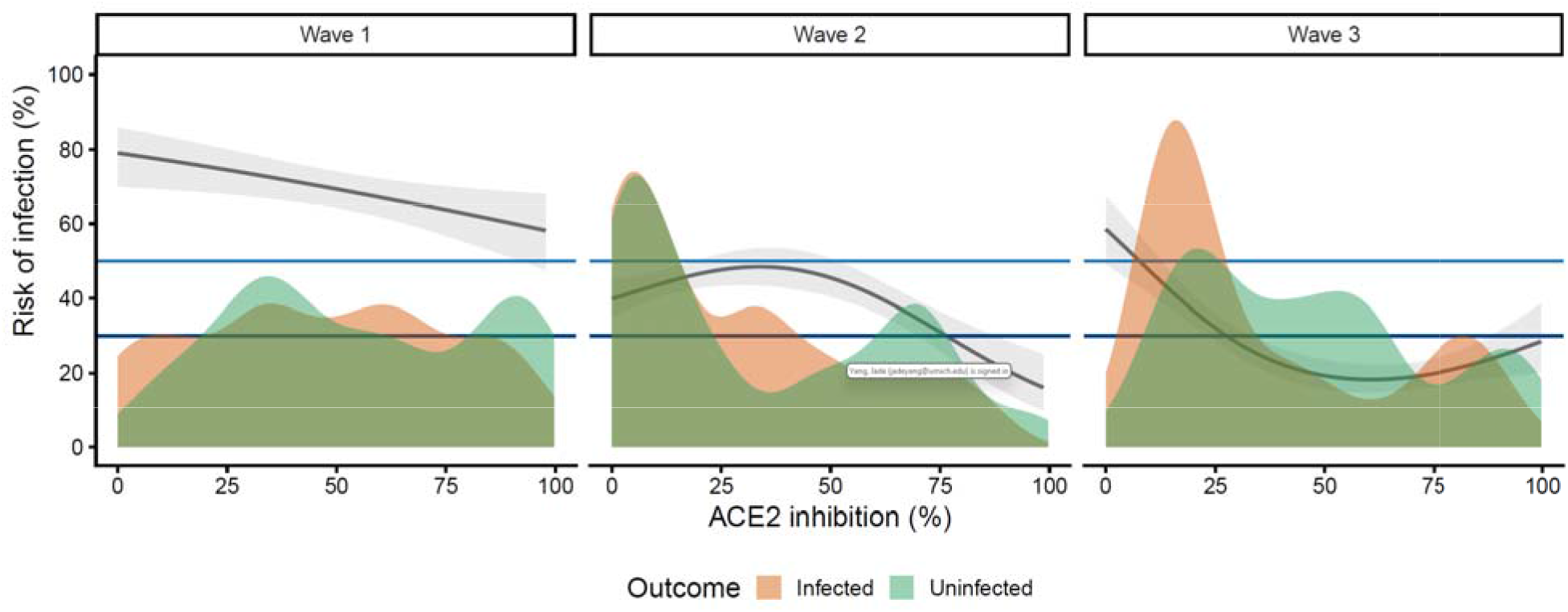
Antibody levels as a correlate of protection during multiple omicron waves, HIVE cohort (2021—2025). Panels show the association between anti-S IgG antibody concentration (A) and ACE2 percent inhibition (B), and corresponding wave-specific analyses (C, D), with the predicted probability of infection. The solid lines represent the mean predicted infection risk based on generalized additive mixed models (GAMMs) fitted to the observed data (see Methods). Shaded ribbons show 95% confidence intervals (CIs) around the predicted infection estimates. Horizontal reference lines indicate the 30% and 50% infection thresholds, corresponding to the antibody levels required to achieve 70% and 50% protection, defined as (1-risk of infected %)*100, respectively. Density plot embedded within the panels display the distribution of observed antibody levels, stratified by infection outcome (infected in orange vs. not infected in green). Each serum specimen initiated an infection risk interval, with the measured antibody level assigned to that interval until infection, censoring, or the next serum collection; participants could contribute multiple intervals over follow-up. IgG concentrations and ACE2 inhibition were measured using the MSD multiplex platform, with composite measures calculated based on antigens representing the dominant circulating SARS-CoV-2 strains during each period.

In age-adjusted models, prior SARS-CoV-2 infection was associated with a lower hazard of symptomatic infection (HR = 0.58, 95% CI: 0.45–0.73, p<0.001) compared to no prior infection, and recent COVID-19 vaccination within the previous 180 days was not associated with significant difference in infection hazard (HR = 1.07, 95% CI: 0.93–1.25, p=0.34) as compared to those who did not receive vaccination in past 180 days (Table 3).

We next assessed whether prior infection and recent vaccination remained associated with a lower infection hazard after adjusting for antibody levels. Prior infection remained associated with a lower hazard of infection (HR = 0.57, 95% CI: 0.45–0.72, p<0.001), whereas recent vaccination was not associated with a lower infection hazard (HR=1.01, 95% CI: 0.88-1.16, p=0.88) (Tables S9-S10).

### 3.5 ACE2 percent inhibition and infection hazard during Omicron waves

Higher ACE2 percent inhibition was also associated with a reduced hazard of symptomatic infection. A 10% increase in ACE2 inhibition corresponded to a hazard ratio of 0.93 (95% CI: 0.90–0.95, p<0.001) (Table 3). After including both symptomatic and asymptomatic infections, each 10% increase in ACE2 inhibition corresponded to a hazard ratio of 0.93 (95% CI: 0.90–0.95, p<0.001) (Table S11).

Across the observed range of ACE2 inhibition values, increasing inhibition was associated with a higher probability of protection. Among our study participants, GAMM-predicted infection risk decreased as ACE2 inhibition increased. The ACE2 inhibition levels corresponding to predicted infection probabilities of 50% and 30%, respectively (equivalent to 50% and 70% predicted protection), were approximately 10.5% and 99.7% (Figure 4). The peak ACE2 inhibition predicted by the immunogenicity model (72.6%) was associated with an estimated infection probability of 35.3% (95% CI: 32.3%–38.3%), corresponding to approximately 64.7% predicted protection against all infections.

We next assessed whether prior infection and recent vaccination remained associated with infection hazard after adjusting for ACE2 inhibition. Prior infection remained protective (HR = 0.52, 95% CI: 0.41–0.66, p<0.001), whereas recent vaccination was not associated with a significant difference in infection hazard (HR = 1.10, 95% CI: 0.95–1.23, p=0.39) as compared to no recent vaccination (Table S11-S12).

## Discussion

Using data from the HIVE longitudinal household cohort, we evaluated antibody correlates of protection against symptomatic SARS-CoV-2 Omicron infections from July 2021 through January 2025. We found that higher IgG concentrations were strongly associated with reduced risk of infection. Specifically, each two-fold increase in IgG concentration was associated with a 15% lower risk of infection. In the subset of 33 participants under weekly surveillance with nasal swab collection, allowing detection of both symptomatic and asymptomatic infections, the association between IgG concentration and infection risk remained statistically significant. Similar patterns were observed in models using ACE2 percent inhibition as the antibody measure.

Our analysis leveraged the unique features of the HIVE longitudinal cohort to evaluate antibody correlates of protection across the Omicron period in children and adults continually followed since before the emergence of Omicron. By modeling infections occurring over multiple Omicron waves within a unified framework using a composite antibody variable, we were able to assess protection across changing variant circulation rather than restricting analyses to a single epidemic period. The cohort’s detailed longitudinal data on vaccination, prior infections, and symptomatic infection histories allowed us to account for previous exposures when evaluating infection risk. In addition, a subset of participants under weekly surveillance enabled the detection of asymptomatic infections, and results from this subgroup were consistent with those from symptomatic-only analyses, supporting the robustness of our findings. However, only a small number of asymptomatic infections were identified, limiting our ability to evaluate correlates of protection against asymptomatic infection separately that the overall estimates were driven primarily by symptomatic infections. Antibody responses were measured using both binding IgG concentrations and functional ACE2 inhibition derived from the same multiplex assay platform, providing complementary measures of immune response while ensuring consistency in laboratory methods.

These findings are consistent with results from clinical trials and other observational studies^22–26^, demonstrating that higher SARS-CoV-2 binding and functional antibody responses are associated with lower risk of symptomatic infection^24,25^. Similar to previous reports, we observed the relationship between increasing antibody levels and decreasing infection risk. However, the antibody concentrations associated with protection in our study were higher than those reported in pre-Omicron and early Omicron wave studies, likely reflecting the increased immune escape of Omicron variants. Differences in study design may also have contributed, as our analyses incorporated multiple serologic measurements over multiple years rather than relying on a single baseline antibody measurement, although antibody waning between serum collection dates was not directly measured.

Our findings also highlight the protection from prior infection and vaccination. After adjusting for antibody concentrations, prior SARS-CoV-2 infection remained significantly associated with reduced risk of subsequent infection, suggesting that infection-induced protection extends beyond that captured by circulating antibody levels alone and may provide broader protection against reinfection, consistent with previous studies^27–29^. This may reflect other immune mechanisms that contribute to protection, including mucosal and tissue-resident immune responses in the respiratory tract, as well as memory B-cell responses, mucosal immunity, and T-cell mediated immunity, which may provide protection even when antibody levels decline^30–32^. In contrast, recent vaccination was not directly associated with reduced infection risk after accounting for antibody levels, suggesting that COVID-19 vaccines predominantly offer protection via antibody induction. Our immunogenicity analyses showed that vaccination was associated with substantially increased IgG concentrations and ACE2 inhibition. However, given the limited sample size, these findings should not be interpreted as establishing vaccine protection through immune responses not measured in this study.

We also identified evidence of an antibody “ceiling effect” following repeated vaccination^29^. One possible explanation is that pre-vaccination antibody concentrations remained relatively high before subsequent doses, resulting in smaller relative increases and an apparent plateau after three to four doses. This is consistent with studies documenting SARS-CoV-2 antibody waning over time^33–35^, followed by boosting to high levels after receiving subsequent exposure. In our data, binding IgG concentrations plateaued after repeated vaccination; however, ACE2 inhibition continued to rise at a slower rate, even after the fourth dose. Because ACE2 inhibition has a similar mechanism of neutralization assays, reflecting the functional ability of antibodies to block viral entry, these findings suggest that while binding antibody levels may plateau, functional activity continues to improve with additional antigenic exposures. This distinction highlights the importance of measuring both binding and functional antibody responses to fully characterize the immune landscape after repeated vaccination. However, our immunogenicity analyses were based on the cumulative number of vaccine doses and did not explicitly account for the interval between vaccine doses or between vaccination and prior infection. Therefore, differences in exposure timing and antibody waning may have contributed to the observed plateauing pattern.

Several limitations should be considered. First, the relatively small and geographically limited sample may limit generalizability, and residual confounding may remain. Second, COVID-19 vaccination was uncommon among children in our cohort, resulting in limited ability to evaluate vaccine-associated antibody responses and protection in younger age groups. Although vaccination histories were collected longitudinally, interim vaccinations may not have been completely captured, potentially contributing to underascertainment. Third, asymptomatic infections could only be systematically detected among participants undergoing weekly swab surveillance. Therefore, asymptomatic infections may have been missed among participants followed through ARI surveillance, potentially leading to misclassification of prior infection history. The small number of detected asymptomatic infections also limited our ability to evaluate correlates of protection against asymptomatic infection separately.

Our findings highlight the critical role of antibody levels (generated from prior infection and vaccination) in reducing infection risk during the Omicron period, and an additional level of protection conferred by prior infection beyond the antibody levels generated. Our findings also suggest functional benefits of booster vaccination despite a possible ceiling in IgG concentrations.

## Author Contributions

Conceptualization: Y.Y., E.T.M

Lab testing: C.J., E.J, W.J.F

Project Administration: A.C.

Data Curation: A.C., M.S., E.G., Y.Y.

Analysis: Y.Y.

Visualization: Y.Y., A.C.

Original draft writing: Y.Y.

Review and editing: A.C., M.S., C.J., E.G., E.J., A.S.L, W.J.F, C.M.M., J.M.J., A.S.M., E.T.M

Funding acquisition: E.T.M., A.S.M.

Supervision: E.T.M.

## Disclaimer

The findings and conclusions in this report are those of the authors and do not necessarily represent the official position of the US Centers for Disease Control and Prevention, the National Institute of Allergy and Infectious Diseases, or the National Institutes of Health.

## Ethics Approval Statement

The University of Michigan Institutional Review Board gave ethical approval for this work.

## Patient consent statement

All patients consented to participate in the study.

## Acknowledgements

We would like to thank all individuals for their participation in the HIVE study. Virus sequencing and analysis in the Lauring laboratory was supported in part by U19 AI181767.

## Data and code availability

All data associated with the study will be available upon request. All original code is deposited at GitHub: link to add and is publicly available.

